## Supplementary Figures 1-4 for "Sensitive detection of SARS-CoV-2 seroconversion by flow cytometry reveals the presence of nucleoprotein-reactive antibodies in unexposed individuals"

**SUPPLEMENTAL INFORMATION**

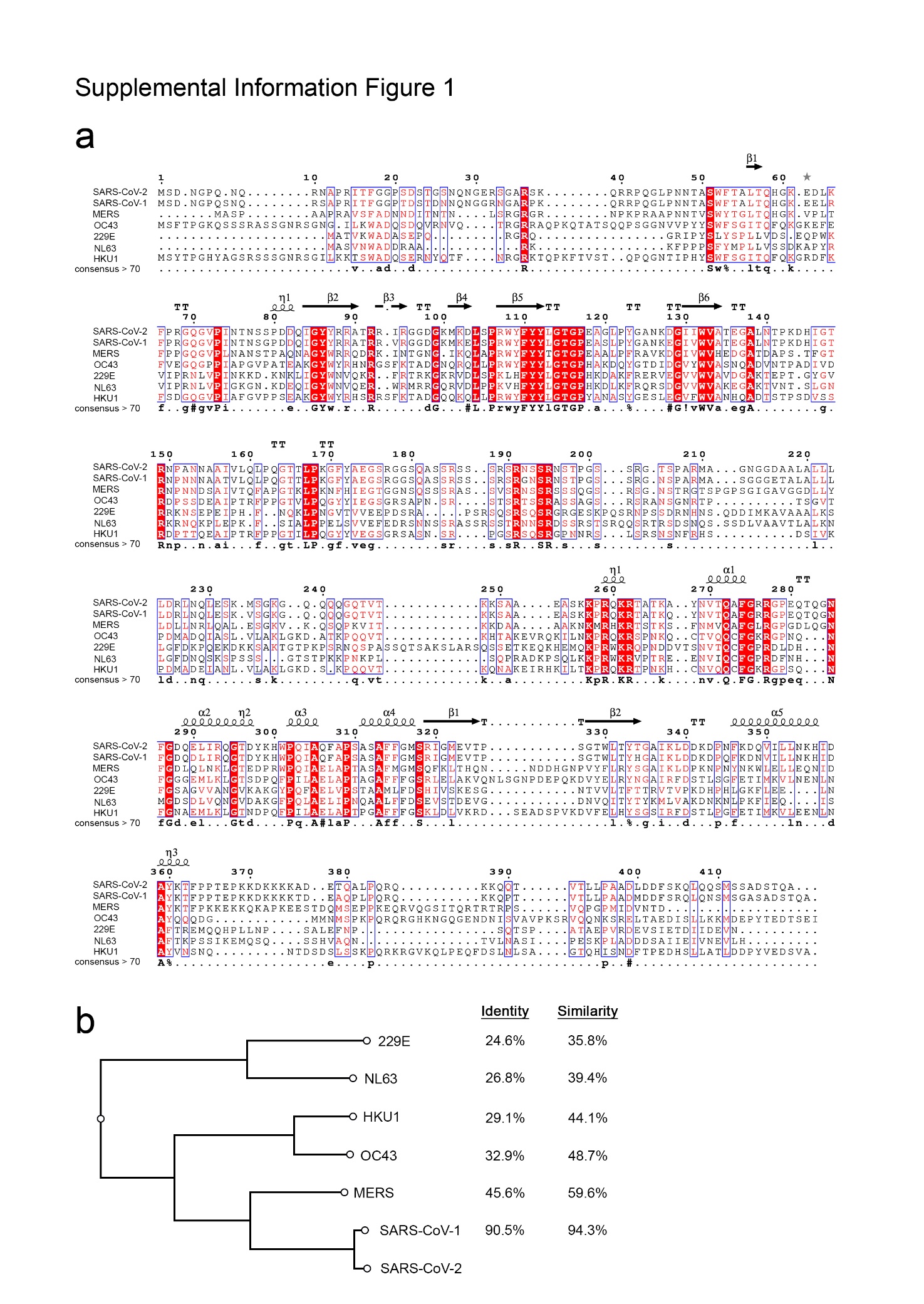

**Supplementary Figure 1.** **Conservation of the N protein sequence across different members of the Coronaviridae family**. **(A)** Alignment of the sequences (Uniprot codes: SARS-CoV-2, P0DTC9; SARS-CoV-1, P59595; MERS-CoV-1, R9UM87; OC43, P33469; 229E, P15130; NL63, Q6Q1R8; and UKU1, Q5MQC6) using Clustal Omega and represented using ESPRIPT 3.0 software using the 70% of equivalent residues calculated per columns, considering physico-chemical properties; on top of the sequences the secondary structural elements derived from the 3D structures determined for the RNA-binding domain (PDB ID: VYO) and the C-terminal dimerization domain (PDB ID: 6WJI) and corresponding to the most conserved regions. **(B)** Phylogram generated from the FASTA alignment file using FastTree. The two columns on the right represent the sequence identity and similarity in a pairwise alignment with the SARS-CoV-2.

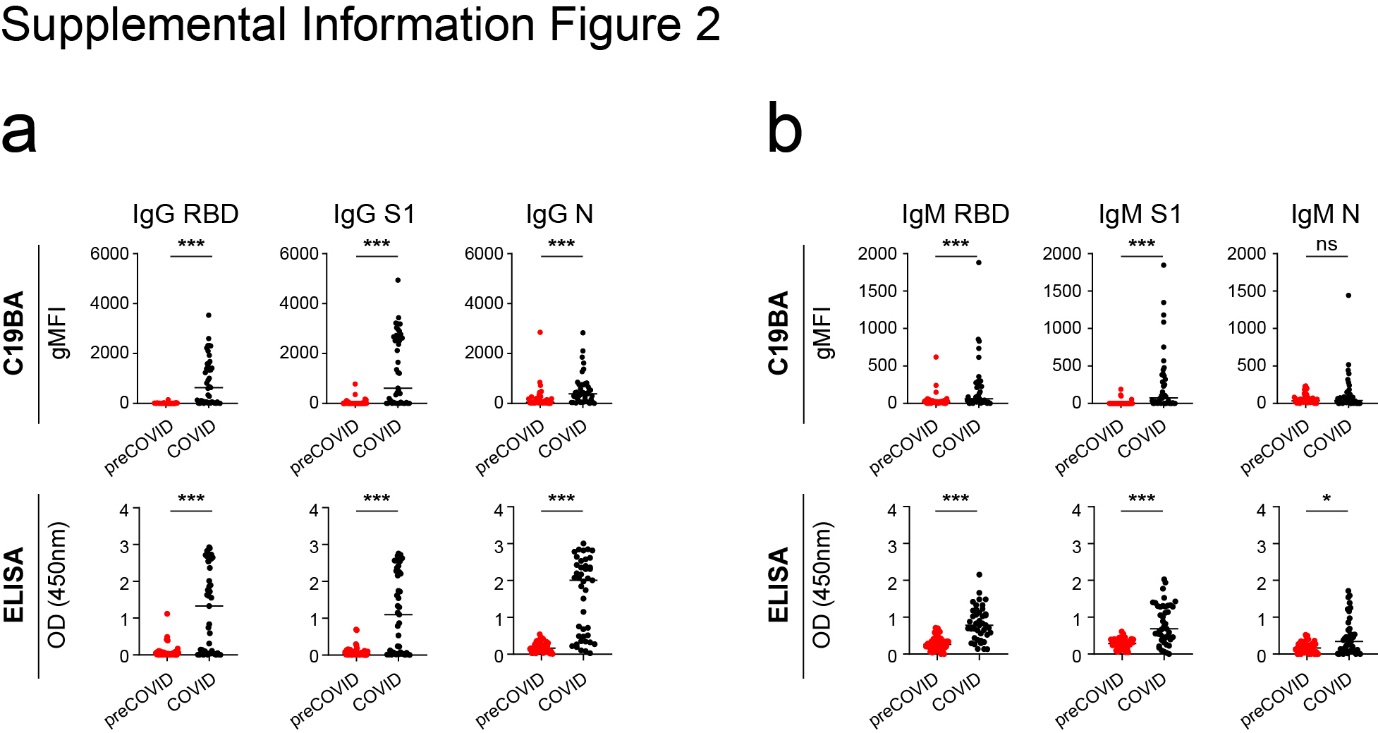

**Supplementary Figure 2.** **Serological responses against RBD, S1 and N measured by C19BA and ELISA.** **(A, B)** gMFI values obtained by C19BA (top) and OD values obtained by ELISA (bottom) for the same samples for IgG (A) and IgM (B). Statistical analyses were performed using an unpaired two-tailed Student’s t-test to compare the preCOVID (red, n=50) and COVID (black, n=43). Asterisks represent p values (*** p<0.001, * p<0.05, ns: not significant). Horizontal lines represent median values.

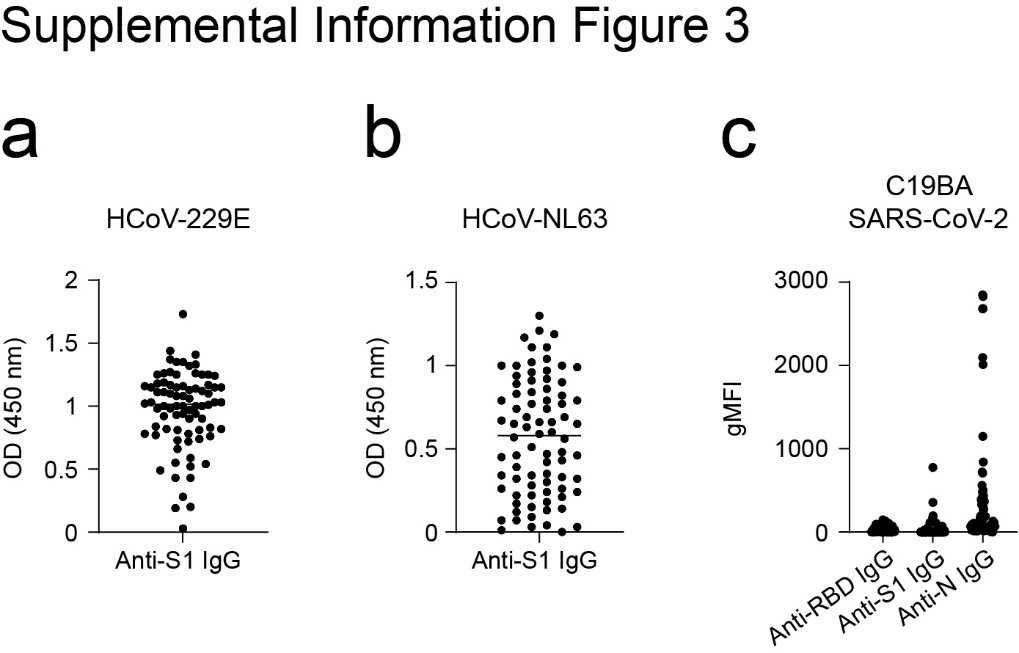

**Supplementary Figure 3. C19BA performed on seropositive samples against common cold coronavirus reveals that RBD, but not N, is specific for determining SARS-CoV-2 seroconversion. (A)** Determination of the presence of IgG antibodies against the spike of common coronavirus HCoV-229E (left) or HCoV-NL63 (right) on serum samples from Covid-negative individuals.**(B)** Levels of IgG antibodies against the indicated SARS-CoV-2 antigens measured by C19BA on the samples shown in A.

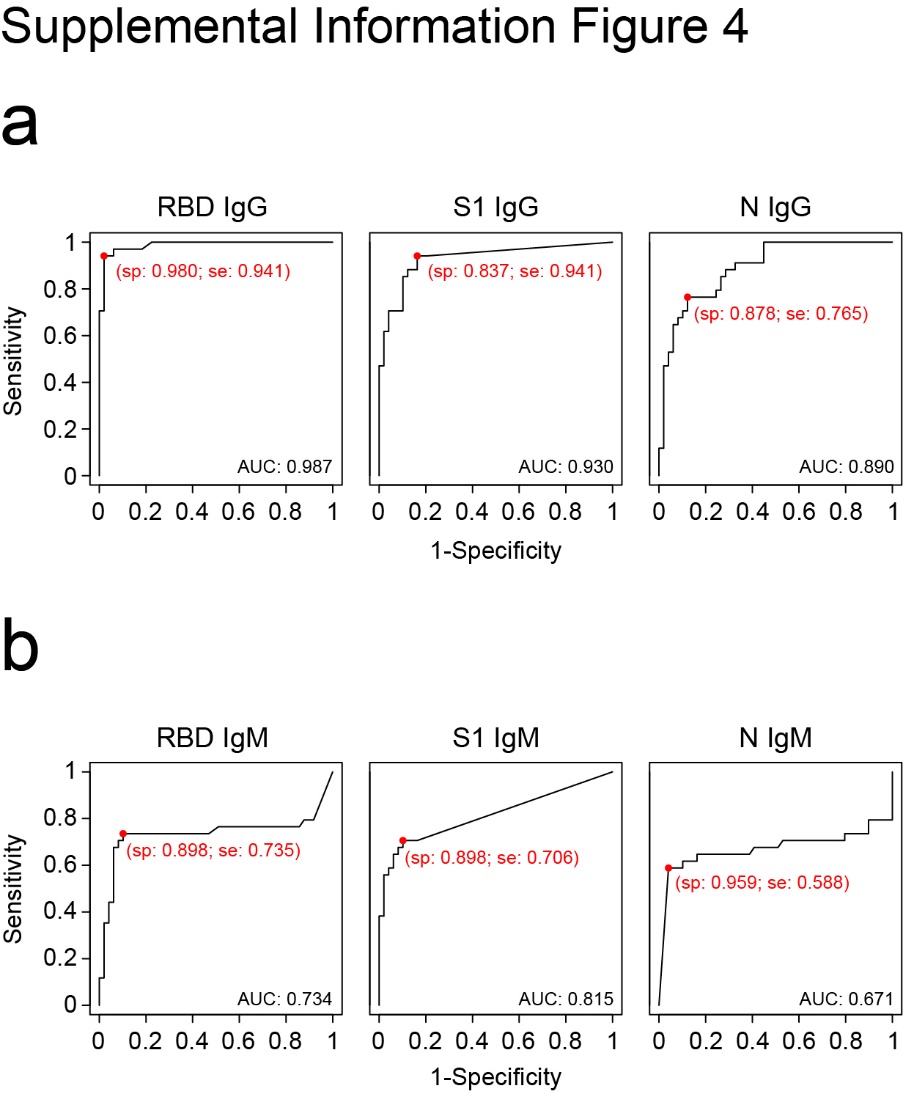

**Supplementary Figure 4.** ROC curves calculated from data obtained by the C19BA assay corresponding to IgG and IgM antibodies against RBD, S1 or N, from preCOVID (n=50) or COVID samples (n=34). Specificity (sp) and sensitivity (se) values for the selected cut-offs are shown in red. AUC: Area Under the Curve.
